# A stop-gain variant in *BTNL9* is associated with atherogenic lipid profiles

**DOI:** 10.1101/2022.06.22.22276448

**Authors:** Jenna C. Carlson, Mohanraj Krishnan, Samantha L. Rosenthal, Emily M. Russell, Jerry Z. Zhang, Nicola L. Hawley, Jaye Moors, Hong Cheng, Nicola Dalbeth, Janak R. de Zoysa, Huti Watson, Muhammad Qasim, Rinki Murphy, Take Naseri, Muagututi’a Sefuiva Reupena, Satupa‘itea Viali, Lisa K. Stamp, John Tuitele, Erin E. Kershaw, Ranjan Deka, Stephen T. McGarvey, Tony R. Merriman, Daniel E. Weeks, Ryan L. Minster.

**Affiliations:** Department of Biostatistics, University of Pittsburgh, Pittsburgh, Pennsylvania, United States of America; Department of Human Genetics, University of Pittsburgh, Pittsburgh, Pennsylvania, United States of America; Center for Craniofacial and Dental Genetics, Department of Oral Biology, University of Pittsburgh, Pittsburgh, Pennsylvania, United States of America; Department of Chronic Disease Epidemiology, Yale School of Public Health, New Haven, Connecticut, United States of America; Department of Biochemistry, University of Otago, Dunedin, New Zealand; Department of Environmental Health, College of Medicine, University of Cincinnati, Cincinnati, Ohio, United States of America; Department of Medicine, Faculty of Medical and Health Sciences, University of Auckland, Auckland, New Zealand; Ngāti Porou Hauora Charitable Trust, Te Puia Springs, Tairāwhiti East Coast, New Zealand; Maurice Wilkins Centre for Molecular Biodiscovery, University of Auckland, Auckland, New Zealand; Ministry of Health, Government of Samoa, Apia, Samoa; Lutia i Puava ae Mapu i Fagalele, Apia, Samoa; School of Medicine, National University of Samoa, Apia, Samoa; Department of Medicine, University of Otago Christchurch, Christchurch, New Zealand; Department of Public Health, Government of American Samoa, Pago Pago, American Samoa; Division of Endocrinology, Department of Medicine, University of Pittsburgh, Pittsburgh, Pennsylvania, United States of America; International Health Institute, Department of Epidemiology, Brown University, Providence, Rhode Island, United States of America; Department of Anthropology, Brown University, Providence, Rhode Island, United States of America

## Abstract

Current understanding of lipid genetics has come mainly from studies in European-ancestry populations; limited effort has focused on Polynesian populations, whose unique population history and high prevalence of dyslipidemia may provide insight into the biological foundations of variation in lipid levels. Here we performed an association study to fine map a suggestive association on 5q35 with high-density lipoprotein cholesterol (HDL-C) seen in Micronesian and Polynesian populations. Fine-mapping analyses in a cohort of 2,851 Samoan adults highlighted an association between a stop-gain variant (rs200884524; c.652C>T, p.R218*; posterior probability = 0.9987) in *BTNL9* and both lower HDL-C and greater triglycerides (TG). Meta-analysis across this and several other cohorts of Polynesian ancestry from Samoa, American Samoa, and Aotearoa New Zealand confirmed the presence of this association (β_HDL-C_ = -1.60 mg/dL, *p*_HDL-C_ = 7.63 × 10^−10^; β_TG_ = 12.00 mg/dL, *p*_TG_ = 3.82 × 10^−7^). While this variant appears to be Polynesian-specific, there is also evidence of association from other multi-ancestry analyses in this region. This work provides evidence of a previously unexplored contributor to the genetic architecture of lipid levels and underscores the importance of genetic analyses in understudied populations.

## INTRODUCTION

Atherogenic lipid profiles – increased total cholesterol (TC), low-density lipoprotein cholesterol (LDL-C), and triglycerides (TG) as well as decreased high-density lipoprotein cholesterol (HDL-C) – are well-documented and heritable risk factors for cardiovascular disease (CVD) worldwide. While behavioral modifications and medication have been successful in improving lipid profiles, CVD is still the leading cause of death worldwide, particularly among people of Polynesian and Pacific Island ancestry (1–5).

The examination of the genetic underpinnings of lipid variation through genome-wide association studies (GWAS) has identified numerous genetic associations. These discoveries have furthered the understanding of cardiovascular disease and potential therapeutic targets; however, this research has primarily come from studies in European-ancestry populations. Recent efforts to diversify research on this topic have highlighted the importance of including diverse populations for gene discovery, yielding novel associations, improved fine-mapping, and better polygenic risk scores (6). Despite this, limited effort has focused on Polynesian populations, whose unique population history including genetic drift from founder effects, small population sizes, and population bottlenecks may provide insight into the biological foundations of variation in lipid levels, which would not only benefit Polynesian individuals, but also those from other populations (7–9).

One region of interest is 5q35, which has previously been associated with HDL-C levels in Micronesian and Samoan populations (10,11). The causal variant at this locus, and the biological mechanism underlying this association with an atherogenic lipid profile is unknown.

The aim of this study was to fine map this association signal in a cohort of 2,851 Samoan adults and replicate this association in several independent Polynesian cohorts from Samoa, American Samoa, and Aotearoa New Zealand. We identified a strong candidate causal variant at this locus, rs200884524 – a missense variant in *BTNL9* – that is associated with lower HDL-C and higher TG concentrations.

## SUBJECTS AND METHODS

### Discovery

We performed association testing between variants on 5q35 and lipid levels in a discovery cohort comprising 2,851 Samoan adults (at least 18 years-old) drawn from a population-based sample recruited from Samoa in 2010 (Table 1), data for which are available from dbGaP (accession number: phs000914.v1.p1). The sample selection, data collection methods, and phenotyping, including the laboratory assays for serum lipid and lipoprotein levels, have been previously described (12,13). Briefly, serum lipid levels (TC, HDL-C, LDL-C, and TG) were derived from fasting whole blood samples collected after a minimum 10-hour overnight fast. LDL-C levels were estimated using the Friedewald equation (14), which does not accurately estimate LDL-C in individuals with TG ≥ 400 mg/dL, and therefore, those individuals missing LDL-C values. Analyses were performed without consideration of hypolipidemic medication, as it was not measured. However, individuals who reported taking medication for heart disease were excluded from analyses, as previous sensitivity analyses identified an association between self-reported use of heart disease medication and TC and LDL-C (10).

**Table 1.** Characteristics of discovery and replication cohorts. Mean (SD) are given for age, total cholesterol (TC), high-density lipoprotein cholesterol (HDL-C), low-density lipoprotein cholesterol (LDL-C), and triglycerides (TG) for each cohort. When applicable, the number of participants missing the measurement (*N miss*) is given.

|  | Samoa Discovery |  | Samoa/American Samoa Replication |  |  |  |  |  |  |  | Aotearoa New Zealand Replication |  |  |  |  |  |
| --- | --- | --- | --- | --- | --- | --- | --- | --- | --- | --- | --- | --- | --- | --- | --- | --- |
|  | n=2,851 |  | n=1,466 |  |  |  |  |  |  |  | n=1,810 |  |  |  |  |  |
|  |  |  | 1994-95 Recruitment (n=557) |  |  |  | 2002-03 Recruitment (n=909) |  |  |  |  |  |  |  |  |  |
|  |  |  | Samoa |  | American Samoa |  | Samoa |  | American Samoa |  | East Polynesia (n=1,109) |  | West Polynesia (n=603) |  | East/West Polynesia (n=98) |  |
|  | Female | Male | Female | Male | Female | Male | Female | Male | Female | Male | Female | Male | Female | Male | Female | Male |
|  | n=1,705 | n=1,146 | n=202 | n=184 | n=96 | n=75 | n=195 | n=185 | n=308 | n=221 | n=456 | n=653 | n=182 | n=421 | n=38 | n=60 |
| **Age**<br>(years) |  |  |  |  |  |  |  |  |  |  |  |  |  |  |  |  |
| Mean | 44.77 | 45.56 | 43.5 | 43.08 | 42.53 | 45.4 | 45.05 | 42.01 | 42.96 | 43.24 | 51.23 | 52.72 | 44.05 | 44.46 | 37.66 | 39.48 |
| SD | 11.09 | 11.29 | 8.49 | 8.81 | 7.57 | 10.5 | 16.79 | 15.72 | 16.03 | 16.42 | 15.48 | 14.32 | 15.71 | 13.99 | 13.77 | 16.15 |
| **TC**<br>(mg/dL) |  |  |  |  |  |  |  |  |  |  |  |  |  |  |  |  |
| Mean | 199.23 | 200.27 | 199.35 | 196.79 | 198.68 | 194.57 | 203.87 | 195.86 | 187.39 | 188.71 | 191.41 | 191.55 | 180.42 | 190.12 | 178.9 | 193.74 |
| SD | 36.13 | 38.66 | 36.88 | 35.07 | 30.63 | 30.46 | 35.92 | 38.05 | 38.97 | 37.17 | 42.36 | 45.54 | 45.41 | 43.61 | 43.01 | 40.53 |
| N Miss |  |  |  |  |  |  |  |  |  |  | 1 |  | 1 |  |  |  |
| **HDL-C**<br>(mg/dL) |  |  |  |  |  |  |  |  |  |  |  |  |  |  |  |  |
| Mean | 46.48 | 43.7 | 43.87 | 41.38 | 34.75 | 32.39 | 47.46 | 45.7 | 42 | 38.54 | 49.27 | 42.36 | 47.8 | 41.42 | 52.66 | 41.46 |
| SD | 10.82 | 11.18 | 10.62 | 11.02 | 7.66 | 8.22 | 10.37 | 11.48 | 8.25 | 8.86 | 15.08 | 12.52 | 12.79 | 12.32 | 14.18 | 12.17 |
| N Miss |  |  |  | 3 |  | 4 | 1 |  |  |  | 1 |  | 1 | 1 |  |  |
| **LDL-C**<br>(mg/dL) <sup>a</sup> |  |  |  |  |  |  |  |  |  |  |  |  |  |  |  |  |
| Mean | 130.17 | 130.2 | 139.12 | 137.45 | 141.34 | 134.57 | 134.65 | 127.27 | 118.85 | 117.48 | 107.67 | 110.29 | 100.6 | 113.79 | 95.85 | 119.37 |
| SD | 32.43 | 34.38 | 33.22 | 32.37 | 30.85 | 35.91 | 32.39 | 36.57 | 34.58 | 33.57 | 34.82 | 40.94 | 37.18 | 39.16 | 35.7 | 32.63 |
| N Miss | 10 | 25 | 1 | 5 |  | 9 | 1 | 3 | 4 | 19 | 34 | 76 | 9 | 37 | 2 | 8 |
| **TG**<br>(mg/dL) |  |  |  |  |  |  |  |  |  |  |  |  |  |  |  |  |
| Mean | 114.97 | 139.16 | 85.01 | 101.42 | 115.2 | 172.83 | 109.38 | 119.26 | 131.28 | 191.64 | 180.88 | 216.8 | 163.07 | 201.47 | 145.98 | 208.49 |
| SD | 80.47 | 112.7 | 76.41 | 95.75 | 52.17 | 121.92 | 54.4 | 94.9 | 75.68 | 171.68 | 120.22 | 153.45 | 117.71 | 133.27 | 94.24 | 137.91 |

<sup>a</sup>LDL-C was estimated using the Friedewald equation so individuals with TG ≥ 400 mg/dL have missing LDL-C values.

This study was approved by the institutional review board of Brown University and the Health Research Committee of the Samoa Ministry of Health. All participants gave written informed consent via consent forms in the Samoan language.

Genotyping was performed using Genome-Wide Human SNP 6.0 arrays (Affymetrix) and extensive quality control was conducted via a pipeline developed by Laurie et al. (15). Additional details for sample genotyping and genotype quality control are described in Minster et al. (13). A Samoan-specific genotype reference panel for imputation was created using the whole-genome sequence (WGS) data from the Trans-Omics for Precision Medicine (TOPMed) program from the National Institutes of Health’s Heart, Lung, and Blood Institute (NHLBI). The reference panel is comprised of high-quality (i.e., passing all QC filters and with a minimum depth of 10), bi-allelic markers from the freeze 8 call subset of the 1,284 individuals from the Samoan Adiposity Study. The reference panel was phased using Eagle v2.4.1 (16). The Samoan-specific reference panel was then used to impute genotype data for remaining Samoan participants in the discovery cohort who were not in the reference panel. Briefly, genotyped variant coordinates were converted to the hg38 genome build using LiftOver (https://genome.ucsc.edu/cgi-bin/hgLiftOver). Then, data were aligned to the reference panel using Genotype Harmonizer (17), using the mafAlign option to align to the minor allele when LD alignment failed and the minor allele frequency (MAF) was ≤ 30% in both the input and reference sets. The resulting variants were then phased with Eagle v2.4.1 (16) and genome-wide imputation was performed using Minimac4 (18). Imputed genotype dosages from variants passing QC (*r*^2^ > 0.3) were combined with genotyped variants for association testing.

We performed preliminary association testing between variants in a 1 Mb region around the sentinel SNP from the preliminary GWAS (10) (hg38 chr5:180414895-181414895) and the four lipid levels (TC, HDL-C, LDL-C, and TG) in the discovery cohort using linear mixed modeling with inverse-normally transformed traits, marginally rescaled variance (to restore it to the original variance before the transformation), and additive genotype coding as implemented in the *GENESIS* R package (19,20) adjusting for the following fixed effects: three principal components of ancestry (generated through PC-AiR (21)), age, age^2^, sex, age × sex interaction, and age^2^ × sex interaction. Age was mean centered for all analyses to avoid multicollinearity issues. Relatedness was measured through an empirical kinship matrix generated with *GENESIS* and was modeled through a random effect. Then, to estimate the effect size of rs200884524 for use in the meta-analysis across cohorts, we used a linear mixed modeling strategy on the original lipid levels (i.e., not transformed) adjusting for the same fixed and random effects as implemented in the *lmekin* function in the *coxme* R package (22). These models were also repeated for several cardiometabolic traits as described in Minster et al. 2016 (13) to examine the potential pleiotropic effects of rs200884524.

As a sensitivity analysis to check for robustness against deviations from normality, the association tests with rs200884524 were repeated using lipid level residuals that were adjusted for three principal components of ancestry, age, age^2^, sex, and age × sex and age^2^ × sex interactions and were then inverse-normally transformed (23) using *RankNorm* function in the *RNOmni* R package (24). To preserve the interpretation of the effect estimates, the results of the untransformed trait analyses are presented. The results of the sensitivity analysis were similar to the models in which lipid levels were not transformed (Table S1).

To detect a potential secondary signal, conditional analyses were run in the 5q35 region in the discovery cohort. rs200884524, the lead SNP in the region, was modeled as a fixed-effect covariate; the rest of the model parameters stayed the same.

Bayesian fine mapping of the discovery cohort results was performed using PAINTOR v2.1 (25). Annotations of variant impact (HIGH, MODERATE, LOW/MODIFIER) as indicated by VEP v91.1 (26) were used to inform the fine mapping.

Evidence of selection at this locus was assessed using *nS*_L_ (number of segregation sites by length) analysis in 419 unrelated Samoans from the discovery cohort with no evidence of non-Oceanic admixture (27). Whole-genome sequencing was completed by TOPMed and a MAF of >0.01 was used. Haplotype phasing was completed using Eagle v2.4.1 (16). *nS*_L_ was calculated with *selscan* in R (28).

### Replication

#### Samoan/American Samoan Replication Cohorts

Two cohorts consisting of a total of 1,466 Samoan and American Samoan adults (at least 18 years-old) were used for replication. The first was a longitudinal study of adiposity and cardiovascular disease risk factors among adults from American Samoa and Samoa recruited between 1994 and 1995, during which lipid levels were measured from fasting serum samples. LDL-C levels were estimated using the Friedewald equation (14), which does not accurately estimate LDL-C in individuals with TG ≥ 400 mg/dL, and therefore, those individuals missing LDL-C values. Detailed descriptions of the sampling, recruitment, and phenotyping have been reported previously (29–31). The second study included adults from American Samoa and Samoa, recruited in 2002–2003, and was drawn from an extended family-based genetic linkage study of cardiometabolic traits (32–34). Probands and relatives were unselected for obesity or related phenotypes, and all individuals self-reported Samoan ancestry. The recruitment process, criteria used for inclusion in this study, and phenotyping have been described in detail previously (35,36). To account for temporal differences, each of the two studies was modeled separately – Samoa/American Samoa 1994-95 (*n* = 557) and Samoa/American Samoa 2002-03 (*n* = 909). Participants in these two cohorts were genotyped using the Infinium Global Screening Array-24 v3.0 BeadChip (Illumina, CA, USA) with custom content that included rs200884524. These studies were approved by the institutional review boards of Brown University and the American Samoa Department of Health, as well as the Health Research Committee of the Samoa Ministry of Health. All participants gave written informed consent.

#### Aotearoa New Zealand Replication Cohorts

A total of 1,810 adults (at least 16 years-old) of Polynesian (NZ Maori and Pacific) ancestry were recruited from Aotearoa New Zealand as part of a study of risk factors for gout, type 2 diabetes, and kidney disease. Lipid parameters (VLDL and TG) have previously been associated with gout in an Aotearoa NZ Polynesian cohort (37). Participants were divided into three replication cohorts based on the self-reported ancestry of their grandparents (East Polynesia [Aotearoa New Zealand Māori and Cook Island Māori] *n* = 1,109; West Polynesia [Samoa, Tonga, Pukapuka, and Niue], *n* = 603; and Mixed East/West Polynesia, *n* = 98). A separate Māori sample set from the rohe (area) of the Ngāti Porou iwi (tribe) of the Tairāwhiti (East Coast of the North Island of New Zealand) region was also included in the East Polynesia group. This sample set was recruited in collaboration with the Ngāti Porou Hauora (Health Service) Charitable Trust. The details of sampling, recruitment, phenotyping, and creation of principal components of ancestry have been previously reported (38,39). Briefly, lipid levels were measured from serum samples at Southern Community Laboratories (Dunedin, NZ). LDL-C levels were estimated using the Friedewald equation (14), which does not accurately estimate LDL-C in individuals with TG ≥ 400 mg/dL, and therefore, those individuals missing LDL-C values. Genotyping of rs200884524 was carried out using the TaqMan® SNP genotyping assay technology (Applied Biosystems, Foster City, USA) using a LightCycler® 480 Real-Time Polymerase Chain Reaction (PCR) system (Roche, Indianapolis, USA). This study was approved by the New Zealand Multi-Region Ethics Committee and the Northern Y Region Health Research Ethics Committee. All participants gave written informed consent.

#### Replication Analyses

Association testing for rs200884524, the sentinel SNP in the 5q35 region as identified in the discovery cohort, and the lipid levels (TC, HDL-C, LDL-C, and TG) was performed in each of the five replication cohorts using linear mixed modeling, adjusting for the following fixed effects: principal components (PCs) of ancestry (four PCs generated through PC-AiR (21) were used for the Samoan/American Samoan replication cohorts, three PCs for the Aotearoa New Zealand replication cohorts), polity (an indicator of Samoa/American Samoa in the two respective cohorts), age, age^2^, sex, age × sex interaction, and age^2^ × sex interaction. Age was mean centered within each cohort for all analyses to avoid multicollinearity issues. Relatedness was measured through empirical kinship matrices and modeled as a random effect using the *lmekin* function in the *coxme* R package (22). For the Samoan/American Samoan replication cohorts, the empirical kinship matrix was generated with *GENESIS* (19,20); for the Aotearoa New Zealand replication cohorts, the empirical kinship matrix was calculated in PLINK v1.9 (40,41) as described previously (38).

### Meta-analysis

The effects of rs200884524 on the four lipid traits from the discovery and 5 replication cohort analyses were combined using an inverse-variance fixed-effect meta-analysis as implemented in the *rmeta* R package (42). Additionally, meta-analyses were conducted separately across the two Samoan/American Samoan replication cohorts and the three Aotearoa New Zealand replication cohorts. Heterogeneity was assessed with Cochran’s Q statistic.

## RESULTS

Descriptions of age, TC, HDL-C, LDL-C, and TG levels in the discovery and replication cohorts are given in Table 1. Generally, the cohorts were very similar (with the exception of TG which varies more widely across cohorts).

The association analyses in the discovery cohort in the 1 Mb region on 5q35 (Figures S1-4) highlighted an association between a stop-gain variant (rs200884524; c.652C>T, p.R218*) in the Butyrophilin Like 9 (*BTNL9*) gene with HDL-C (*p* = 4.05 × 10^−8^; Figure S1). Conditional analyses on rs200884524 showed some evidence of a secondary signal approximately 0.449 Mb upstream from the lead variant in the region (rs71680280; CA>C, conditional *p* = 6.52 × 10^−6^, LD with rs200884524 *r*^2^ = 0.035, Figure S1). Bayesian fine mapping pointed to rs200884524 with high posterior probability of causality (PP = 0.9987); all other variants in the region had posterior probability < 0.10. rs200884524 did not show evidence of association with any other cardiometabolic phenotype measured (Table S1).

The estimated effects of rs200884524 on the four lipid levels for each cohort are given in Table 2. rs200884524 was significantly associated with lower average HDL-C levels overall (meta-analysis β = -1.60 mg/dL, *p* = 7.63 × 10^−10^; Figure 1), with no evidence of heterogeneity of effect across cohorts (*p* = 0.65). Similarly, rs200884524 was significantly associated with higher average TG levels overall (meta-analysis β = 12.00 mg/dL, *p* = 3.82 × 10^−7^, heterogeneity *p* = 0.75). Sensitivity analyses using inverse-normal-transformed phenotypes showed similar results (Table S2). rs200884524 accounts for 0.94% and 0.68% of the variation in HDL-C and TG respectively in the discovery cohort.

**Table 2.** rs200884524 association results for untransformed lipid levels: total cholesterol (TC), HDL cholesterol (HDL-C), LDL cholesterol (LDL-C), and triglycerides (TG). For individual cohorts, βs, 95% confidence intervals (CI), and *p*-values obtained from linear mixed models adjusting for fixed effects of principal components of ancestry, polity (Samoa/American Samoa), age, age^2^, sex, age × sex interaction, and age^2^ × sex interaction. Age was mean centered for all analyses to avoid multicollinearity issues. Relatedness was measured through an empirical kinship matrix and was modeled through a random effect. For meta-analyses, βs, 95% confidence intervals (CI), and *p*-values were obtained from inverse-variance fixed-effect meta-analyses. Heterogeneity *p*-values are from Cochran’s Q test. Results with *p*-values < 0.05 are indicated in bold.

|  | TC | HDL-C | LDL-C | TG |
| --- | --- | --- | --- | --- |
| | $\beta$ (95% CI) mg/dL | $\beta$ (95% CI) mg/dL | $\beta$ (95% CI) mg/dL | $\beta$ (95% CI) mg/dL |
| Cohort | $p$-value | $p$-value | $p$-value | $p$-value |
| **Samoa Discovery (n=2,851; MAF = 0.223)** | -1.65 (-3.91, 0.61) | **-1.84 (-2.54, -1.14)** | -0.93 (-2.98, 1.12) | **13.48 (7.49, 19.47)** |
|  | 0.15 | **$2.74 \times 10^{-7}$** | 0.37 | **$1.04 \times 10^{-5}$** |
| **Samoa/American Samoan Replication Cohorts** |  |  |  |  |
| 1994-95 Samoan/American Samoan (n=557; MAF = 0.233) | 0.68 (-3.91, 5.26) | -1.98 (-3.35, -0.60) | 0.85 (-3.64, 5.34) | 15.60 (3.67, 27.53) |
| 2002-03 Samoa/American Samoan (n=909; MAF = 0.202) | 0.76 (-3.27, 4.79) | -0.83 (-1.99, 0.33) | 0.71 (-3.09, 4.50) | 4.65 (-7.88, 17.17) |
| *Meta Analysis of 2 Samoan/American Samoan Replication Cohorts* | 0.72 (-2.30, 3.75) | **-1.31 (-2.19, -0.42)** | 0.77 (-2.13, 3.66) | **10.39 (1.75, 19.03)** |
| *Meta Analysis Effect p-value* | 0.64 | **$3.86 \times 10^{-3}$** | 0.60 | **0.018** |
| *Heterogeneity p-value* | 0.98 | 0.21 | 0.96 | 0.21 |
| **Aotearoa New Zealand Replication Cohorts** |  |  |  |  |
| Eastern Polynesian (n=1,109; MAF = 0.049) | -1.40 (-9.57, 6.76) | -0.58 (-3.16, 1.99) | -1.63 (-9.10, 5.84) | 15.52 (-10.85, 41.88) |
| Western Polynesian (n=603; MAF = 0.216) | -1.21 (-6.98, 4.56) | -1.54 (-3.21, 0.14) | 0.06 (-5.27, 5.40) | 4.90 (-11.97, 21.76) |
| Mixed Eastern/Western Polynesian (n=98; MAF = 0.133) | 0.48 (-15.27, 16.24) | -2.90 (-7.71, 1.91) | 4.88 (-8.05, 17.80) | 12.41 (-35.63, 60.44) |
| *Meta Analysis of 3 Aotearoa New Zealand Replication Cohorts* | -1.13 (-5.65, 3.39) | **-1.38 (-2.73, -0.04)** | 0.04 (-4.08, 4.15) | 8.34 (-5.29, 21.96) |
| *Meta Analysis Effect p-value* | 0.62 | **0.044** | 0.99 | 0.23 |
| *Heterogeneity p-value* | 0.98 | 0.68 | 0.69 | 0.79 |
| **Meta Analysis (all cohorts)** | -0.85 (-2.53, 0.83) | **-1.60 (-2.11, -1.09)** | -0.31 (-1.86, 1.24) | **12.00 (7.37, 16.63)** |
| *Meta Analysis Effect p-value* | 0.32 | **$7.63 \times 10^{-10}$** | 0.70 | **$3.82 \times 10^{-7}$** |
| *Heterogeneity p-value* | 0.90 | 0.65 | 0.89 | 0.75 |

**Figure 1.**
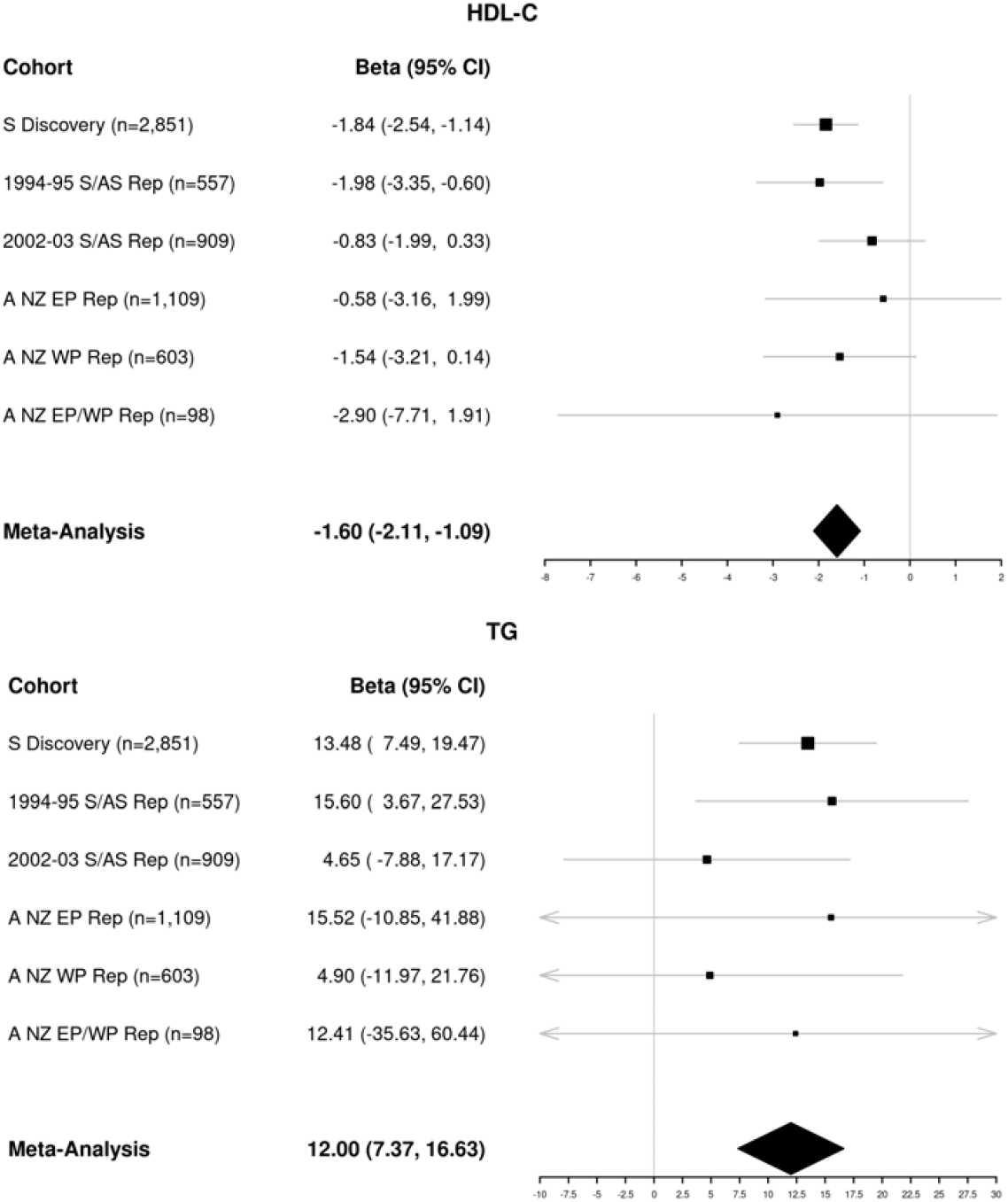
Meta-analysis results of rs200884524 for high-density lipoprotein cholesterol (HDL-C) and triglycerides (TG). Effect estimates (Beta) and 95% confidence intervals (CI) for HDL-C (mg/dL) and TG (mg/dL) are given for each cohort (abbreviations: Samoan [S], American Samoan [AS], Replication [Rep], Aotearoa New Zealand [A NZ], Eastern Polynesian [EP], Western Polynesian [WP]). Meta analysis results were obtained using inverse-variance fixed-effects meta-analysis. Arrows indicate confidence intervals that have been truncated for plotting.

rs200884524 has MAF <0.0001 in gnomAD v3.1.1 (accessed 16 Feb 2022); of the 14 alleles seen in gnomAD, 7 are from East Asians, with no observed homozygotes. However, we observed a MAF of 0.223 in the Samoan discovery cohort with 163 observed homozygotes. The MAFs of rs200884524 ranged from 0.049 to 0.233 in the Samoan/American Samoan and Aotearoa New Zealand replication cohorts, with higher frequencies observed among the Western Polynesian participants (Table 2). Consistent with these disparate allele frequencies, rs200884524 also showed evidence of positive selection in the discovery cohort (*nS*_L_ score of 1.79, 98.9 percentile in the Samoan genome).

## DISCUSSION

We found strong evidence of association between a stop-gain variant in *BTNL9*, rs200884524, and atherogenic serum lipid profiles (lower HDL-C, higher TG) across several independent cohorts of Polynesian individuals. Interestingly, this variant appears to be Polynesian specific – common in those of Western Polynesian ancestry (which includes Samoa), rarer in those of Eastern Polynesian ancestry, and absent from other non-Polynesian populations. When compared to effect sizes of other index variants from GWAS of HDL-C (6), the effect size of rs200884524 was moderate-to-large, which is similar to other findings in this population (13,43,44).

Moreover, the identification of rs200884524 was achieved using a population-specific genotype imputation panel, highlighting the importance of such panels when working with isolated populations. Previous work (using the discovery cohort from these results) observed suggestive association between this region and HDL-C (10), but was limited to SNP array data only, rather than the dense data from imputed genotypes. The sparse genotypes in this region did not facilitate accurate fine mapping. In fact, we previously nominated *MGAT1* as the causal gene for this locus, as the stop-gain variant in *BTNL9* was not genotyped. Once the dense information from imputed genotypes was available, it was easy to identify the stop-gain variant rs200884524 as a variant of interest, which led us to include it with the custom content selected for genotyping in the Samoan/American Samoan replication cohorts. These results highlight the necessity of population-specific imputation reference panels and the fruitfulness of gene discovery in isolated populations.

There is evidence that rs200884524 plays a role in nonsense-mediated decay (NMD) efficacy; the variant is located in exon 4 of 11 in *BTNL9* and has strong mRNA degradation predicted (NMDetective score 0.6 (45,46)). While *BTNL9* is not constrained/predicted to be loss-of-function intolerant (pLoF observed/expected = 0.94, gnomAD v2.1.1), the role of the BTNL9 protein in human disease is still unknown.

Despite the T allele of rs200884524 having very low frequency in other populations, the association between variants in this region and lipid levels does not appear to be Polynesian-specific. Results from studies of the UKBiobank and the Global Lipids Genetics Consortium demonstrate evidence of association in this region as well. Specifically, an intron variant in nearby *BTNL3* (∼61 kb away from rs200884524) was significantly associated with HDL-C in a GWAS of ∼459,00 individuals of European Descent (rs138354839, *p*_HDL-C_ = 9.70 × 10^−13^) (47). Additionally, variants in and upstream of *BTNL8* (∼128-169 kb away from rs200884524) were associated with total and HDL-C in a large, multi-ancestry meta-analysis (rs138692142, *p*_HDL-C_ = 8.09 × 10^−17^; rs188238483 *p*_TC_ = 3.39 × 10^−9^) (6). Each of these variants is rare in the Samoan discovery cohort presented here (MAF = 0.009, 0.0003, and 0.00003, respectively).

Furthermore, there were significant associations between pLoF variants in *BTNL9* and both Apolipoprotein A and HDL-C observed in gene-based studies of European-ancestry individuals also from the UKBiobank (48–50). Both associations are driven by a single variant (rs367635312, *p*_ApoA_ = 1.93 × 10^−9^, *p*_HDL-C_ = 1.48 × 10^−7^, MAF = 0.0125 in non-Finnish Europeans). This variant is also rare in the Samoan discovery cohort (MAF = 0.00016).

Interestingly, the region also harbors a ∼55kb common deletion with consequence in *BTNL8* and *BTNL3* (DEL_5_65831 gnomAD SVs v2.1). This deletion is common in those of Asian, American, and European descent, but rare in African and Oceanic (Papuan and Melanesian) populations (51). The frequency of this deletion in Polynesians is unknown, as Polynesians have differing genetic ancestries from Papuans and Melanesians. Functional analysis has shown that this deletion down-regulates *BTNL9* in lymphoblastoid cell lines (51). This deletion is in high linkage disequilibrium (r^2^ >0.80) with 26 SNVs in the UKBiobank (50,52). The SNV in very high LD with the deletion (rs72494581, LD with deletion 0.97) showed minimal evidence of association with HDL-C in the Pan-UKBiobank (*p* = 0.056 in meta-analysis across all populations, *p* = 0.037 in individuals of European descent) (50,53). Because of the low allele frequency of rs200884524 in non-Polynesian populations, it is unknown if the stop-gain variant is in LD with this deletion. Further work is needed to explore the interplay of this deletion with rs200884524 to examine the causal mechanism and resulting biological impact on lipids.

Little is known about the function of *BTNL9* and the mechanism by which it may impact serum lipids; however, it has been previously associated with cardiomyopathy (54–56), pre-eclampsia (57), and various cancers (58,59). *BTNL9* has a cell- and tissue-specific expression pattern – it is most highly expressed in human B cells of adipose tissue, lung, thymus, spleen, and heart (60). Moreover, BTNL9 is localized to the plasma membrane, and binds to immune cell surfaces (61). BTNL9 belongs to the butyrophilin (BTN) family of membrane proteins which is part of a superfamily of immunoglobulin (Ig) receptors (62). Several studies suggest a role of BTN proteins in inflammatory and immunological functions (61,63–72), supported by the shared extracellular characteristics and homology of the BTN proteins with the B7 family (which includes ligands and receptors for T cell activity) (62). *BTNL9* is member of a BTN subfamily along with *BTNL3* and *BTNL8* which all diverged from a common ancestor (62). While *BTNL3* and *BTNL8* are primate specific, *BTNL9* is conserved across several mammalian species, however, the protein structure differs across species. Human BTNL9 has two substitution mutations that result in the loss of the IgC domain that is present in other mammals (62). In mice, the receptor to Btnl9 was shown to be expressed in immune cells, supporting the hypothesized immune function (62,73). Future functional work is needed to identify the biological role of BTNL9 in lipid and lipoprotein metabolism, immune function, and ultimately, cardiovascular (and potentially cancer) risk.

Additional work is necessary to characterize the relationship between variation in *BTNL9* and lipid levels, especially analyzing the impact of the nearby deletion in *BTNL8*/*BTNL3*, as well as any potential interaction between immune cells and lipid transport pathways. However, these findings fine map the suggestive association in 5q35 to a stop-gain variant in *BTNL9*, providing evidence of a new contributor to the genetic architecture of lipids. This work highlights the importance of measuring association in non-European populations as it can provide insight not only into population-specific findings and benefits, but also to cross-population associations which may lead to therapeutic targets able to benefit multiple population groups.

## Data Availability

The discovery cohort data used for this study are available through dbGaP (accession numbers: phs000914.v1.p1 and phs000972.v5.p1). The Samoan/American Samoan and Aotearoa New Zealand replication cohorts' data have not been deposited in a public repository because participants did not give consent for data sharing. Code used for data analysis will be made available upon request.

## SUPPLEMENTAL INFORMATION

Supplemental Information includes four figures and two tables.

## ACKNOWLEDGMENTS

We would like to thank the Samoan participants of the study, local village authorities, and the many Samoan and other field workers over the years. We acknowledge the Samoan Ministry of Health, the Samoa Bureau of Statistics, and the American Samoan Department of Health for their support of this research. We give particular thanks to two research assistants, Melania Selu and Vaimoana Lupematasila, who contributed to the 2010 recruitment and continue to assist us in our work in Samoa. We would also like to thank the Aotearoa New Zealand participants and to thank Aotearoa New Zealand recruiters Ria Akuhata, Jordyn Allan, Nancy Aupouri, Jill Drake, Carol Ford Roddi Laurence, Christopher Franklin, Meaghan House, Gabrielle Sexton, and Fiona Taylor.

Molecular data for the Trans-Omics in Precision Medicine (TOPMed) program was supported by the National Heart, Lung and Blood Institute (NHLBI). Genome Sequencing for NHLBI TOPMed: Samoan (phs000972.v5.p1) was performed at New York Genome Center Genomics (HHSN268201500016C) and Northwest Genomics Center (HHSN268201100037C). Core support including centralized genomic read mapping and genotype calling, along with variant quality metrics and filtering were provided by the TOPMed Informatics Research Center (3R01HL-117626-02S1; contract HHSN268201800002I). Core support including phenotype harmonization, data management, sample-identity QC, and general program coordination were provided by the TOPMed Data Coordinating Center (R01HL-120393; U01HL-120393; contract HHSN268201800001I). We gratefully acknowledge the studies and participants who provided biological samples and data for TOPMed.

This work was funded by the National Institute of Health grants R01HL093093 (STM), R01HL133040 (RLM), R01AG09375 (STM), R01HL52611 (MI Kamboh), R01DK59642 (STM), R01DK55406 (RD), and by the New Zealand Health Research Council. Genotyping was performed in the Core Genotyping Laboratory at the University of Cincinnati, funded by National Institutes of Health grant P30ES006096 (SM Ho).

