## Supplemental Information for "A stop-gain variant in *BTNL9* is associated with atherogenic lipid profiles"

**Figure S1. Regional association plots for HDL cholesterol (HDL-C) in the discovery cohort.**

**(A)** Association results for HDL-C in the discovery cohort obtained via linear mixed modeling with inverse-normally transformed traits, marginally rescaled variance (to restore it to the original variance before the transformation), and additive genotype coding as implemented in the *GENESIS* R package (1,2). Variants are colored based on LD with rs200884524 (purple). **(B)** Association results for HDL-C conditional upon rs200884524 show suggestive evidence of a secondary signal for rs71680280 in the HDL-C scan. Variants are colored based on LD with rs71680280 (purple). All plots were created in LocusZoom (3).

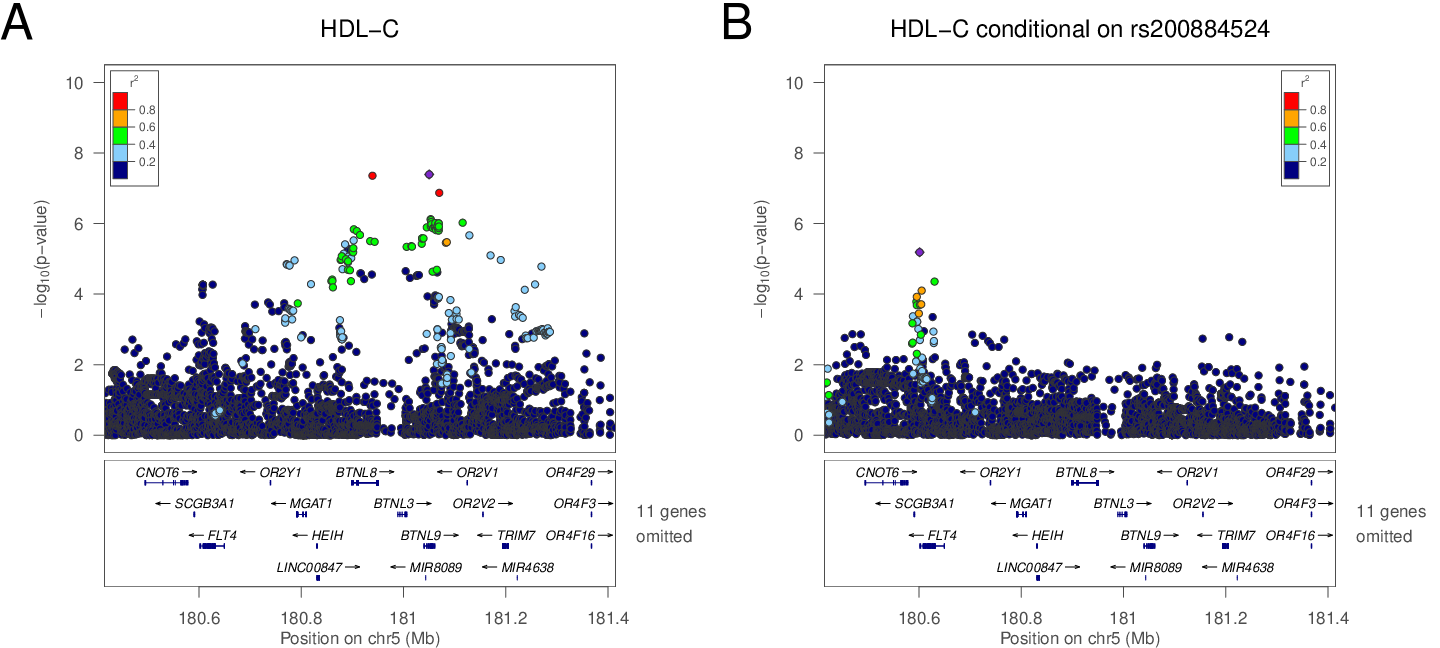

**Figure S2. Regional association plot for triglycerides (TG)** **in the discovery cohort.**

Association results for HDL-C in the discovery cohort obtained via linear mixed modeling with inverse-normally transformed traits, marginally rescaled variance (to restore it to the original variance before the transformation), and additive genotype coding as implemented in the *GENESIS* R package (1,2). Variants are colored based on LD with rs200884524 (purple). All plots were created in LocusZoom (3).

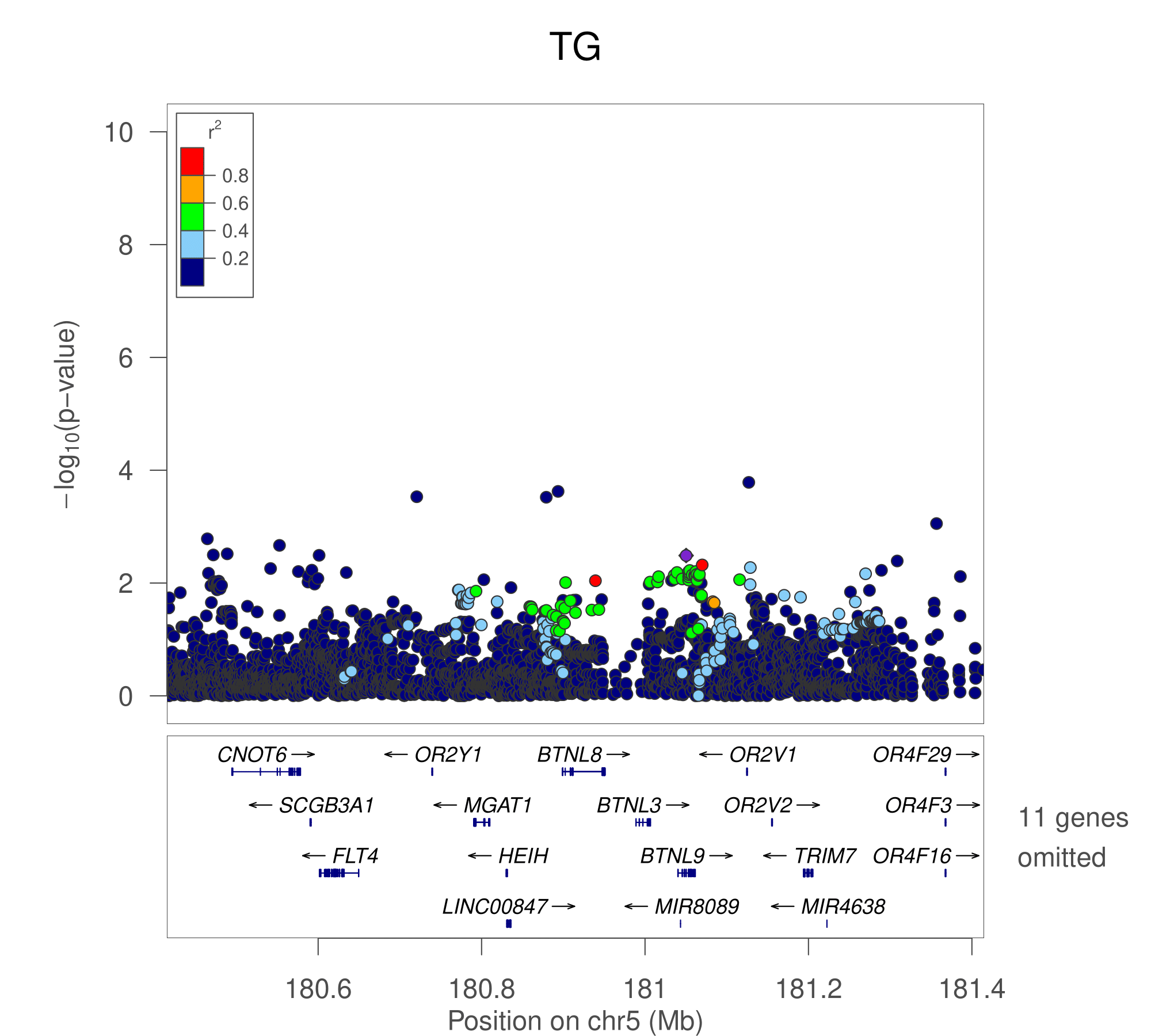

**Figure S3. Regional association plot for LDL cholesterol (LDL-C) in the discovery cohort.**

Association results for HDL-C in the discovery cohort obtained via linear mixed modeling with inverse-normally transformed traits, marginally rescaled variance (to restore it to the original variance before the transformation), and additive genotype coding as implemented in the *GENESIS* R package (1,2). Variants are colored based on LD with rs200884524 (purple). All plots were created in LocusZoom (3).

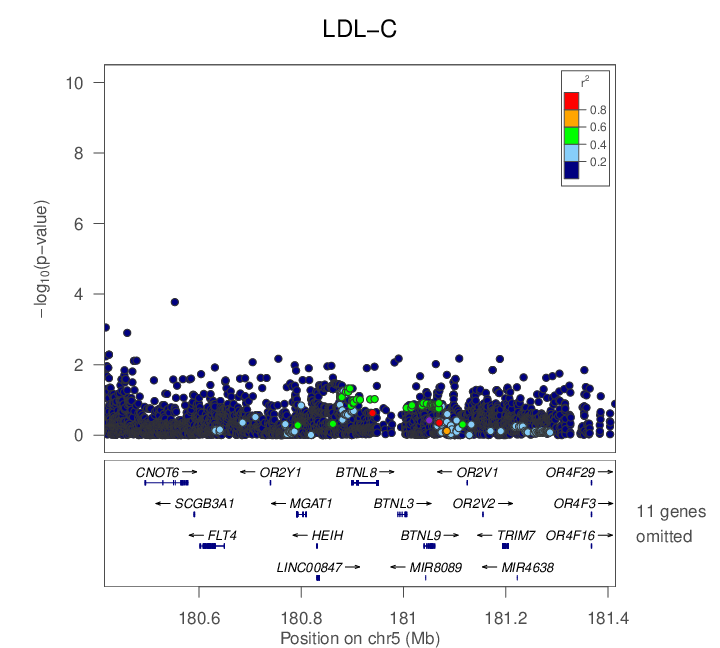

**Figure S4. Regional association plot for total cholesterol (TC) in the discovery cohort.**

Association results for HDL-C in the discovery cohort obtained via linear mixed modeling with inverse-normally transformed traits, marginally rescaled variance (to restore it to the original variance before the transformation), and additive genotype coding as implemented in the *GENESIS* R package (1,2). Variants are colored based on LD with rs200884524 (purple). All plots were created in LocusZoom (3).

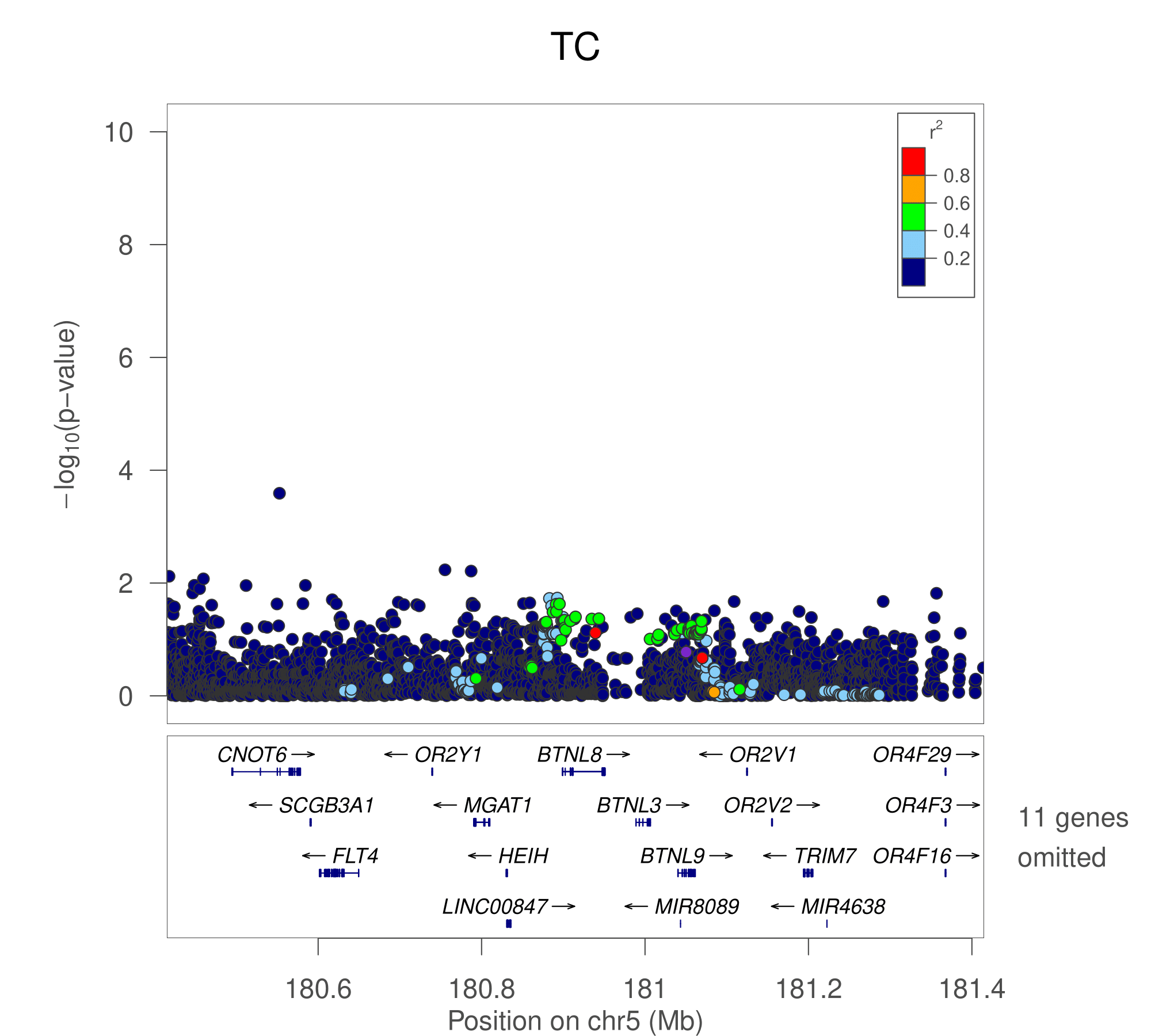

**Table S1. rs200884524 association results for other phenotypes in discovery cohort.** Sample size (N), effect estimate (β) and corresponding standard error (SE), *p*-values, and 95% confidence intervals (CI) were obtained from linear mixed models adjusting for fixed effects of principal components of ancestry, age, age^2^, sex, age × sex interaction, and age^2^ × sex interaction in up to 3,073 Samoan adults from the discovery cohort. Details of the phenotypic measurements are available in Minster et al. 2016 (4). Age was mean centered for all analyses to avoid multicollinearity issues. Relatedness was measured through an empirical kinship matrix and was modeled through a random effect. For leptin, sex-specific models were fit, without adjustment for any terms including sex.

| **Trait** | **N** | **Beta** | **SE** | ***p*-value** | **95% CI** |
| --- | --- | --- | --- | --- | --- |
| **BMI (kg/m2)** | 3073 | 0.14 | 0.20 | 0.50 | (-0.26, 0.54) |
| **Body Fat %** | 2900 | 0.06 | 0.38 | 0.88 | (-0.69, 0.81) |
| **Abdominal Circumference (cm)** | 3064 | 0.71 | 0.45 | 0.12 | (-0.18, 1.59) |
| **Hip Circumference (cm)** | 3065 | 0.33 | 0.37 | 0.38 | (-0.40, 1.06) |
| **Abdominal-hip ratio** | 3063 | 0.003 | 0.002 | 0.07 | (0.00, 0.01) |
| **Fasting glucose (mg/dL)** | 2656 | 0.05 | 1.44 | 0.97 | (-2.76, 2.87) |
| **Fasting insulin (μU/mL)** | 2655 | -0.14 | 0.57 | 0.80 | (-1.26, 0.98) |
| **HOMA-IR** | 2432 | -0.02 | 0.14 | 0.89 | (-0.28, 0.25) |
| **Adiponectin (μg/mL)** | 2864 | -0.06 | 0.09 | 0.50 | (-0.24, 0.12) |
| **Leptin in men (ng/mL)** | 1153 | 0.12 | 0.35 | 0.73 | (-0.57, 0.82) |
| **Leptin in women (ng/mL)** | 1711 | 0.56 | 0.59 | 0.33 | (-0.58, 1.71) |

**Table S2. rs200884524 association results for inverse-normally-transformed lipid levels: total cholesterol (TC), HDL cholesterol (HDL-C), LDL cholesterol (LDL-C), and triglycerides (TG).** For individual cohorts, βs, 95% confidence intervals (CI), and *p*-values obtained from linear mixed models adjusting for fixed effects of principal components of ancestry, polity (Samoa/American Samoa), age, age^2^, sex, age × sex interaction, and age^2^ × sex interaction. Age was mean centered for all analyses to avoid multicollinearity issues. Relatedness was measured through an empirical kinship matrix and was modeled through a random effect. For meta-analyses, βs, 95% confidence intervals (CI), and *p*-values were obtained from inverse-variance fixed-effect meta-analyses. Heterogeneity *p*-values are from Cochran’s Q test. Results with *p*-values < 0.05 are indicated in bold.

|  | **TC ^a^** | **HDL-C ^a^** | **LDL-C ^a^** | **TG ^a^** |
| --- | --- | --- | --- | --- |
|  | **β (95% CI)** | **β (95% CI)** | **β (95% CI)** | **β (95% CI)** |
| **Cohort** | ***p*-value** | ***p*-value** | ***p*-value** | ***p*-value** |
| **Samoan Discovery (n=2,851; MAF = 0.223)** | -0.04 (-0.11, 0.02) | **-0.18 (-0.24, -0.12)** | -0.03 (-0.09, 0.04) | **0.10 (0.03, 0.16)** |
|  | *0.18* | ***3.44 × 10^-8^*** | *0.40* | ***2.78 × 10^-3^*** |
| **Samoan/American Samoan Replication Cohorts** | -0.04 (-0.11, 0.02) | **-0.18 (-0.24, -0.12)** | -0.03 (-0.09, 0.04) | **0.10 (0.03, 0.16)** |
| 1994-95 Samoan/American Samoan (n=557; MAF = 0.233) | 0.02 (-0.12, 0.15) | -0.23 (-0.37, 0.09) | 0.02 (-0.12, 0.17) | 0.12 (-0.02, 0.25) |
| 2002-03 Samoa/American Samoan (n=909; MAF = 0.202) | 0.01 (-0.11, 0.13) | -0.09 (-0.21, 0.03) | 0.02 (-0.11, 0.14) | 0.10 (-0.01, 0.22) |
| *Meta Analysis of 2 Samoan/American Samoan Replication Cohorts* | 0.01 (-0.08, 0.10) | **-0.15 (-0.24, -0.06)** | 0.02 (-0.07, 0.11) | **0.11 (0.02, 0.20)** |
| *Meta Analysis Effect p-value* | *0.77* | ***1.26 × 10^-3^*** | *0.67* | ***0.015*** |
| *Heterogeneity p-value* | *0.97* | *0.13* | *0.95* | *0.85* |
| **Aotearoa New Zealand Replication Cohorts** |  |  |  |  |
| Eastern Polynesian (n=1,109; MAF = 0.049) | -0.04 (-0.22, 0.15) | -0.03 (-0.22, 0.16) | -0.04 (-0.24, 0.15) | 0.06 (-0.13, 0.24) |
| Western Polynesian (n=603; MAF = 0.216) | -0.03 (-0.17, 0.10) | -0.13 (-0.27, 0.00) | 0.00 (-0.14, 0.15) | -0.02 (-0.16, 0.11) |
| Mixed Eastern/Western Polynesian (n=98; MAF = 0.133) | 0.00 (-0.38, 0.39) | -0.24 (-0.63, 0.15) | 0.19 (-0.21, 0.58) | 0.11 (-0.28, 0.50) |
| *Meta Analysis of 3 Aotearoa New Zealand Replication Cohorts* | -0.03 (-0.14, 0.07) | **-0.11 (-0.21, 0.00)** | 0.00 (-0.11, 0.11) | 0.01 ( -0.1, 0.12) |
| *Meta Analysis Effect p-value* | *0.56* | ***0.048*** | *0.97* | *0.85* |
| *Heterogeneity p-value* | *0.98* | 0.52 | *0.59* | *0.69* |
| **Meta Analysis (all cohorts)** | -0.03 (-0.07, 0.02) | **-0.16 (-0.21, -0.11)** | -0.01 (-0.06, 0.04) | **0.08 (0.04, 0.13)** |
| *Meta Analysis Effect p-value* | *0.28* | ***4.52 × 10^-11^*** | *0.70* | ***3.93 × 10^-4^*** |
| *Heterogeneity p-value* | *0.96* | *0.42* | *0.88* | *0.67* |

^a^ trait was inverse-normally transformed
